# Selectively caring for the most severe COVID-19 patients delays ICU bed shortages more than increasing hospital capacity

**DOI:** 10.1101/2020.12.05.20222968

**Authors:** Miles Roberts, Helena Duplechin Seymour, Faculty Advisor, Alexander Dimitrov

## Abstract

SARS-CoV-2, the virus responsible for COVID-19, has killed hundreds of thousands of Americans. Although physical distancing measures played a key role in slowing COVID-19 spread in early 2020, infection rates are now peaking at record levels across the country. Hospitals in several states are threatened with overwhelming numbers of patients, compounding the death toll of COVID-19. Implementing strategies to minimize COVID-19 hospitalizations will be key to controlling the toll of the disease, but non-physical distancing strategies receive relatively little attention. We present a novel system of differential equations designed to predict the relative effects of hospitalizing fewer COVID-19 patients vs increasing ICU bed availability on delaying ICU bed shortages. This model, which we call SEAHIRD, was calibrated to mortality data on two US states with different peak infection times from mid-March – mid-May 2020. It found that hospitalizing fewer COVID-19 patients generally delays ICU bed shortage more than a comparable increase in ICU bed availability. This trend was consistent across both states and across wide ranges of initial conditions and parameter values. We argue that being able to predict which patients will develop severe COVID-19 symptoms, and thus require hospitalization, should be a key objective of future COVID-19 research, as it will allow limited hospital resources to be allocated to individuals that need them most and prevent hospitals from being overwhelmed by COVID-19 cases.

## 2 Introduction

At the time of writing this, John’s Hopkins University reports that about 230,000 Americans have died of COVID-19 (https://coronavirus.jhu.edu/) - the disease caused by the coronavirus SARS-CoV-2. US hospitals are on the front lines in the battle against the rising death toll, especially now as states experience record-high case numbers (https://coronavirus.jhu.edu/). Ever since the initial outbreak, the primary goal of US public health measures has been to slow COVID-19 infection rates enough that hospitals are not overwhelmed by a large influx of diseased individuals, also phrased as “flattening the curve”. This goal has mostly been achieved thus far through physical distancing – any measure that increases the average distance or adds barriers between individuals from different households. Physical distancing is often defined by the closing of workplaces, schools, and non-essential businesses [34], but the wearing of personal protective equipment (PPE) like gloves, face-coverings, and eye protection also match this broad definition. Physical distancing measures have been invaluable for mitigating COVID-19 spread, lowering the growth rate of the disease [9] and preventing at least half a million COVID-19 cases [34]. However, physical distancing is not without drawbacks.

Physical distancing imposes significant economic and psychological costs that are themselves the common subject of COVID-19 models [29]. Closing businesses and restricting travel in response to the COVID-19 pandemic disrupted US supply chains [14], exacerbated social inequalities [11], and increased rates of anxiety and depression [6]. There’s even evidence to suggest that physical or social distancing may increase the spread of conspiracy theories [13]. Furthermore, physical distancing measures are ultimately voluntary and experts expect such measures to be less effective as lockdowns continue and isolated individuals inevitably grow bored [24]. News outlets report that Americans are already experiencing considerable resistance toward continued physical distancing measures, especially the wearing of masks [8]. Physical distancing continues to be the best tool America has for halting COVID-19 spread but considering supplementary strategies may limit the need for physical distancing, and therefore its consequences, in future pandemics.

We present a model that considers two non-physical distancing strategies to prevent overwhelming hospitals with COVID-19 infections: (1) hospitalizing fewer patients with COVID-19 and (2) increasing hospital capacity for COVID-19 patients. The former can be accomplished by selectively hospitalizing only serious COVID-19 cases, allowing less serious cases to self-isolate, while the later can be accomplished by increasing the number of ICU beds, ventilators, PPE, and hospital staff. For simplicity, we focus on just ICU bed counts as a measure of hospital capacity and only consider the states of Washington (WA) and Colorado (CO) because of their similar case numbers but different peak infection times from March - May 2020.

Washington was the first state in the U.S. to have confirmed COVID-19 infections [28]. When COVID-19 first arrived there, it spread rapidly among the elderly and nurses at long-term care facilities [25]. Newspapers at the time reported a reasonable fear that there would not be enough hospital beds in the whole state to care for the coming wave of COVID-19 infected patients [4]. WA enacted stay-at-home orders in mid-March and many other states like CO soon followed, as uncontrolled community-level transmission was already occurring in other states [23]. In fact, early in the pandemic, CO had one of the highest death counts in the US, just behind California [30]. Thanks to policies to expand hospital capacity and enact physical distancing, Washington and CO avoided exceeding the state-wide limit on hospital beds from March - May. However, as of October 28th 2020, John Hopkin’s University reports that WA is experiences 620 new COVID-19 cases every day, up from 370 cases per day in September 2020. Alarmingly, CO is currently reporting over 1700 new COVID-19 cases every day, twice as many as it was reporting in during its last infection peak in late July (https://coronavirus.jhu.edu/data/new-cases-50-states). Thus, knowing the relative benefits of increasing hospital capacity vs decreasing hospitalization in either of these states is still relevant today. Neither of these strategies should replace physical distancing, of course, but they can still contribute to mitigation efforts.

## 3 Methods

We employed a system of ordinary differential equations to model the spread of COVID-19 in WA and CO. We chose to model just WA and CO because of their similar case numbers during their initial outbreaks, their high-quality reporting according to COVID-tracking project (see Table S1), but different peak infection times. We further focused on data from only the period of March 15th – May 18th as this captured the majority of the first “infection wave” in both states. Most of the model parameters were estimated from previous studies on COVID-19 spread. The remaining parameters were estimated by minimizing the sum of squared errors between the model and data on cumulative deaths over time. Our model was calibrated to mortality data only because, due to testing limitations, counts of COVID-19 deaths are generally considered more reliable than counts of COVID-19 cases [3]. All simulations and analyses were conducted in R [31]. Simulation, optimization, and data visualization were all executed with the deSolve, Flexible Modeling Environment (FME), and ggplot2 packages respectively [35],[36],[41].

### 3.1 Gathering Data

All of the datasets input into our model model are listed in Table S2, along with hyperlinks to their associated websites. Counts of COVID-19 deaths during the period of March 15th – May 18th were downloaded from the novel coronavirus infection map hosted by the University of Washington. Counts of on COVID-19 hospitalizations were downloaded from a publicly available database hosted by Definitive Healthcare, a healthcare data analytics company (https://www.definitivehc.com/about). To correct for under-reporting of hospitalizations, data on hospitalization response rates were downloaded from each state’s respective Department of Health (DOH) website. This data has since been removed from the WA DOH website, but a copy of the dataset is included in the supplemental (Table S3). Next, the number of ICU beds in WA was downloaded directly from the WA DOH website, but this information could not be found on the CO DOH website. Thus, a report of 2018 ICU bed counts from a local CO news organization was used instead. Finally, population size estimates for both states were acquired from their respective US Census Bureau websites.

### 3.2 Correction for under-reporting by hospitals

Not all hospitals report counts of COVID-19 cases to their state governments every day. For the states we studied, fewer hospitals reported COVID-19 hospitalizations during weekends than weekdays (see Tables S3 and S4). The lack of daily reporting partly contributes to hospitalization reports underestimating the true number of hospitalizations. To account for under-reporting of hospitalizations, we applied a simple correction based on the assumption that the ratio of hospitalizations to number of hospitals was the same for the sample of reporting hospitals in a state as it is for all hospitals in a state. In other words:

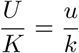

Where U is the number of hospitalizations in the entire state, K is the number of hospitals in the entire state, u is the number of reported hospitalizations, and k is number of reporting hospitals. U is the only unknown in this equation, meaning it can be calculated as:

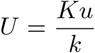

To apply this correction to the entire time series, we wrote U and u as functions of time U(t) and u(t) respectively:

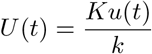

The WA DOH only counted the number of reporting hospitals for May 9th – May 15th, so data from this period was extrapolated to the entire time span of March 15th – May 18th (see Table S3). There are 92 acute care hospitals in the entire state, but only 66 hospitals reported case counts every day for May 9th – May 15th on average. Thus:

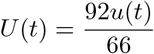

The CO DOH website also reports the percentage of hospitals that update COVID-19 data. However, these data are deleted from the website weekly. Thus, the average percentage of hospitals reporting data had to be estimated from data from July 4th – July 9th (see Table S4). About 75.43 percent of CO hospitals reported COVID-19 cases every day during this period on average. Thus, the applied correction was:

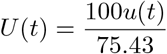

### 3.3 Model formulation

Our system of ordinary differential equations included a total of 7 state variables and 9 parameters, which are defined in Tables S5 and S6 and diagrammed in Figure 1. From Figure 1, the full set of differential equations describing the spread of COVID-19 in our model can be written as:

**Figure 1:**
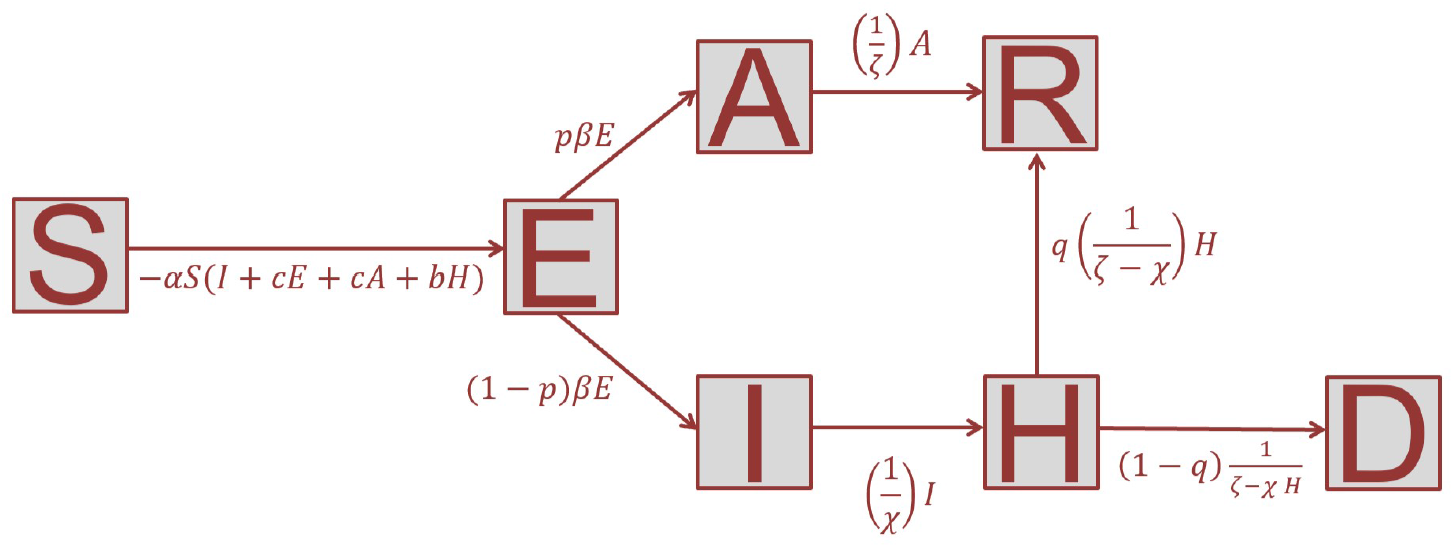
A compartment diagram depicting the system of ordinary differential equations used in this study. See Tables S5 and S6 for variable and parameter definitions.

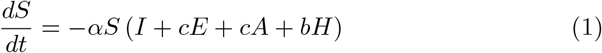

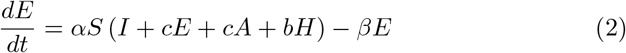

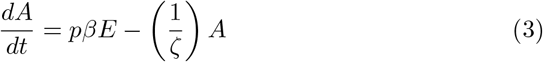

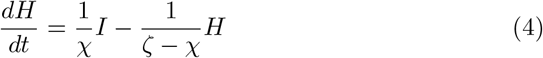

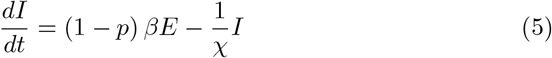

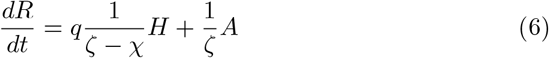

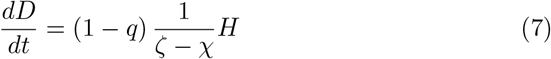

Adhering to epidemiological model naming conventions, this model will be referred to as the SEAHIRD model for the remainder of the paper. The SEAHIRD model begins with assuming that susceptible individuals are exposed to COVID-19 at rates proportional to how often they contact individuals harboring SARS-CoV-2 (*α*).These individuals fall into multiple categories: symptomatic infected (I), exposed but undetermined symptom development (E), asymptomatic infected (A), and symptomatic isolated infected (H, for “home-bound” or “hospitalized”). The scaling factors c and b are meant to reflect the “infectiveness” of individuals in the E, A, and H compartments relative to the I compartment. We chose *c* to be 0.55, reflecting the finding that asymptomatic individuals are 55 percent as infectious as symptomatic individuals [19]. We assumed *b* to be 0.1, reflecting how isolated individuals ideally contact susceptible individuals only rarely.

Once an individual is exposed (E) to SARS-CoV2, they were assumed to have a probability *p* of becoming asymptomatically infected (A) and a probability of 1–*p* of becoming symptomatically infected (I). Furthermore, the rate at which E individuals transformed into A or I individuals was assumed to be proportional the inverse of the incubation period, *β*, estimated as 1/6.6 days [19]. Asymptomatic individuals were assumed to never perish from COVID-19 and to recover from the disease at a rate proportional to 1/*ζ*, the inverse of the recovery period. The recovery period, *ζ*, was estimated from a previous study on COVID-19 severity as 24.7 days [39]. This result agrees with other studies on COVID-19 suggesting that infected asymptomatic individuals typically shed SARS-CoV-2 viruses 15 – 26 days after initial infection [21].

Once an exposed individual (E) individual develops into a symptomatically infected individual (I), they are assumed to mix with the population for *χ* days before isolating themselves (see equations 4 and 5). *χ* was initially assumed to be equal to 1 day in the baseline SEAHIRD models, but we also ran the SEAHIRD model using *χ* values of 0.5 days and 2 days.

Once isolated, the infectiveness of symptomatic individuals was assumed to drop to 0.1*α*. This rate was intentionally made non-zero to account for any minor contact isolated individuals have with susceptible individuals, since quarantine measures are rarely perfect. Once isolated (H), individuals had a probability *q* of recovering from the disease and a probability 1–*q* of dying (see equations 6 and 7). The value of q was estimated as 0.986, again based on COVID-19 studies in China [42]. In addition, some of the rate constants in equations 6 and 7 were assumed to be the inverse of recovery period – *χ*, where the “-*χ*” accounts for the time symptomatic individuals spend mixing with the population before isolating themselves.

For the sake of simplicity, we assume that recovered individuals (R) are not susceptible to re-infection with SARS-CoV-2. This is not known to be strictly true for SARS-CoV-2 infection in humans, but it is reasonable given how non-human primates respond to SARS-CoV-2 infection [7].

### 3.4 Choosing initial conditions

Given that SARS-CoV-2 is a completely novel virus and there is currently no vaccine to invoke immunity, the entire populations of WA and CO were assumed to be susceptible to SARS-CoV-2 infection. Thus, the number of susceptible individuals in either state on March 15th, *S*_0_, was initially estimated as:

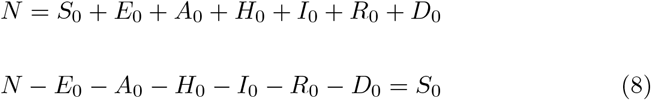

Here, N is the total population size of the state and the other variables are the initial values for the given state’s SEAHIRD compartments. During SEAHIRD model optimization (see section 3.5), *S*_0_ was allowed to vary, but this equation was used to define the optimization algorithm’s parameter search space.

Since COVID-19 mortality data is generally reliable, the number cumulative deaths up until March 15th, *D*_0_, was not adjusted in the raw data. On the other hand, estimating the number of symptomatic, infected individuals required some adjustment. Ideally, the initial number of I individuals (*I*_0_) could be calculated as:

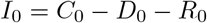

Where *C*_0_, *D*_0_, and *R*_0_ are cumulative numbers of COVID-19 cases, deaths, and recoveries up until March 15th, respectively. For simplicity, *R*_0_ was assumed to be 0, since this category of individuals would ultimately not have much impact on model dynamics, leaving just:

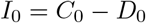

However, *C*_0_ is drastically underestimated for the US population. One preprint study suggests that increasing the number of cases in the US by 279 percent would bring the US death rate down to a similar rate as South Korea, a country with reliable infection counts, after accounting for demographic differences [17]. Thus, *I*_0_ was estimated as:

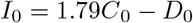

This calculation assumes that the correction for the US applies to individual states as well. The initial number of exposed individuals (*E*_0_) and asymptomatic infected individuals (*A*_0_) was assumed to be directly proportional to *I*_0_. In the absence of reliable data on these numbers, we decided:

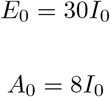

because these numbers generated biologically reasonable model curves and they reflect how the majority of COVID-19 transmission occurs through asymptomatic individuals [19].

Finally, the initial number of symptomatic, isolated individuals (*H*_0_) was assumed to be:

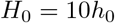

Here, *h*_0_ is the number of individuals hospitalized with COVID-19 on March 15th. In other words, for every individual hospitalized with COVID-19, there were assumed to be nine other symptomatic individuals that were self-isolating at home.

### 3.5 Fitting SEAHIRD to data

We used the Levenberg–Marquardt algorithm implemented in the FME package to optimize *α* and *S*_0_ such that the sum of squared error between the SEAHIRD model output and the curve of cumulative deaths over time for either WA or CO was minimized. In all model fitting cases, the initial estimate of *S*_0_ was given by equation 8 and the initial estimate of *α* was 3E-7. Furthermore, *α* was always bounded between 3E-6 and 3E-8 while *S*_0_ was bounded between 0.05 and 1.5 times its initial estimate. These bounds were chosen because they produced biologically reasonable results (see Figure 2).

**Figure 2:**
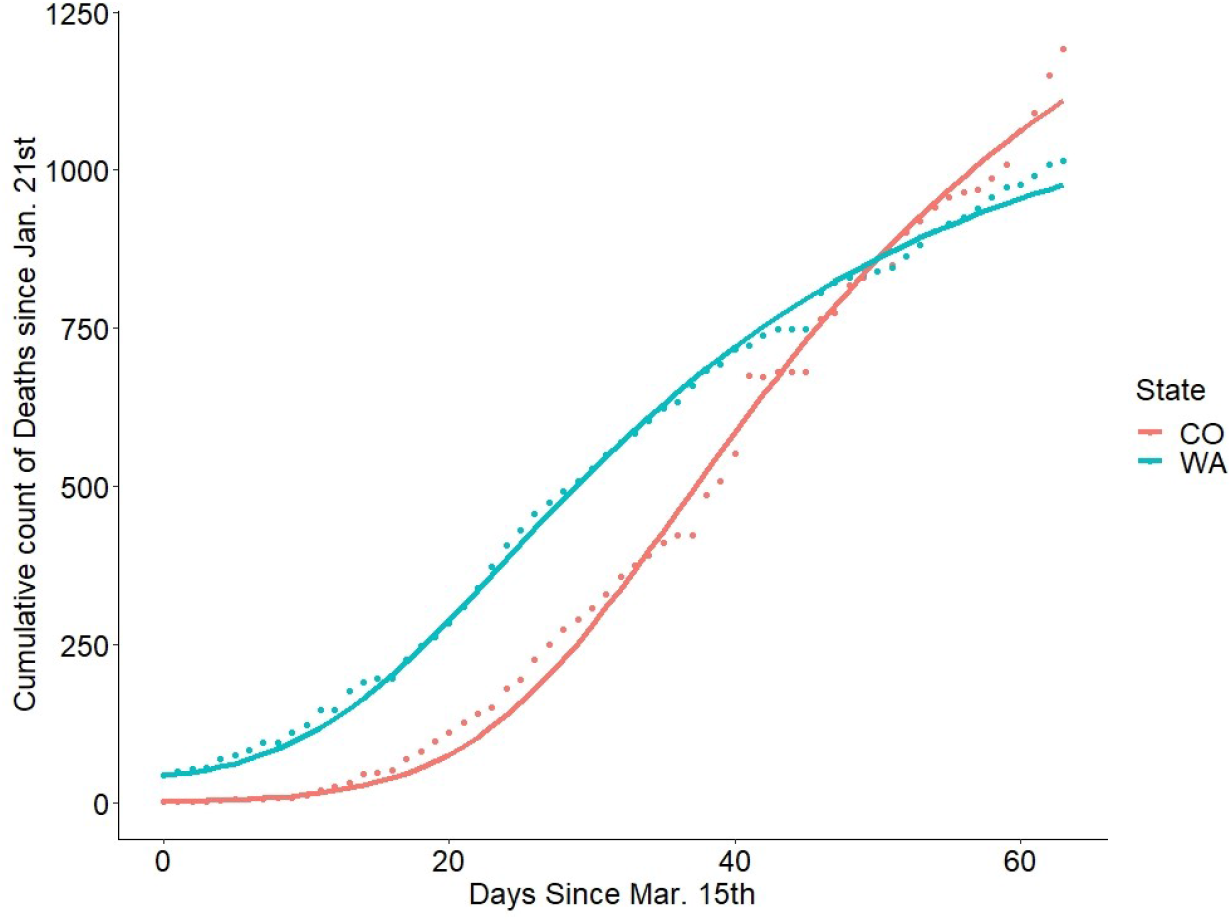
Points are raw counts of deaths due to COVID-19 since Jan. 21^st^ – colored by the state in which the deaths occurred - and the lines are the SEAHIRD model fits to these points. Best fit parameters were *α* = 7.954e-07 and *S*_0_ = 4.893e+05 for the WA SEAHIRD model and *α* = 5.410e-07 and *S*_0_ = 7.477e+05 for the CO SEAHIRD model. For both models *E*_0_ = 30\**I*_0_, *H*_0_ = 10\**h*_0_, and p = 0.86 and *χ* = 1.

### 3.6 Estimating probability of hospitalization given symptomatic infection

The number of individuals hospitalized with COVID-19 was assumed to be a constant fraction of the number of individuals in compartment H. After simulating the SEAHIRD model based on the initial conditions described above, this fraction was estimated numerically by multiplying the H(t) function by 10000 different constants increasing from 0 to 0.1 in increments of 1E-5 and calculating the sum of squared error between the resulting curve and hospitalization counts over time. The constant h that gave the lowest sum of squared error between H(t)*h and the curve of hospitalization counts over time, after correcting for under-reporting, thus estimated the probability of hospitalization given symptomatic infection. This analysis was performed once for the WA dataset and once for CO dataset.

### 3.7 Estimating effect of decreased hospitalization and increased bed cap on delaying bed shortage

Pre-pandemic surveys suggest that about 50 – 80 percent of ICU beds in America are typically occupied at any given time [43]. Thus, we assumed that 30 percent of ICU beds in either WA or CO could be reasonably allocated to COVID-19 infected patients during the modeling period, which is referred to as the hospital “bed cap” at some points in the paper. Once the probability of hospitalization given symptomatic infection (hereto referred to as the “hospitalization probability”) was estimated from the fit of SEAHIRD model to the hospitalization data, we decreased this probability by 50 percent and observed the change in when the bed cap was exceeded under the same SEAHIRD model. Holding this probability at its original estimated value, we then increased the bed cap by 50 percent and recorded the change in when the bed cap was exceeded. Finally, we tested the effect of altering the probability of hospitalization and the bed cap simultaneously by plugging 10000 different combinations of these two numbers spanning values from 0 – 1, in increments of 1E-5, into the SEAHIRD model (see Figures S5 and S9).

### 3.8 Sensitivity analysis

Once we developed the baseline SEAHIRD models (*E*_0_ = 30\**I*_0_, *H*_0_ = 10\**h*_0_, p = 0.86, and *χ* = 1) for CO and WA conditions and parameter values were chosen, we tested the sensitivity of the WA and CO SEAHIRD models in three ways. First, we used the sensFun() command in the FME package to analyze the local sensitivity of the SEAHIRD model to its initial conditions and parameter *α* according to the methods described in the documentation [35]. This analysis was repeated twice, once for WA initial conditions and once for CO initial conditions. Second, we then focused on *E*_0_ and *H*_0_ for both SEAHIRD models and re-ran both models under nine different combinations of *E*_0_ and *H*_0_ values while holding all other initial conditions at their baseline values (see Tables S7 and S8). For each set of initial conditions, we recorded the number of days until 30 percent ICU bed capacity was exceeded, and the number of days until the ICU bed capacity was exceeded if the hospitalization probability was decreased by 50 percent or if statewide ICU bed capacity was increased by 50 percent. Finally, estimates of p vary widely in published literature, so we ran the SEAHIRD model with two different values of p (0.86 and 0.425) while holding all other parameters and conditions at their baseline values. The first value of p comes from a study that estimated the fraction of cases that went undocumented during a period of COVID-19 spread in China [19]. Cases often go undocumented when infected individuals have symptoms that are too mild for them to bother getting tested. Thus, we reasoned this fraction should be close to the value of p. The second value of p from a census in Italy where the authors directly observed 42.5 percent of their COVID-19 infected participants lacking symptoms [18]. Finally, we varied *χ* between three different values (1, 0.5, and 2) while holding all of the other parameters at their baseline and recorded the resulting change in SEAHIRD predictions.

## 4 Results

### 4.1 Effective population sizes for COVID-19 transmission

Figure 2 presents the optimized baseline SEAHIRD model’s fits to mortality data for CO and WA (initial conditions *E*_0_ = 30\**I*_0_, *H*_0_ = 10\**h*_0_, p = 0.86, and *χ* = 1). The best fit parameters for *α* and *S*_0_ in under these conditions were 7.954e-07 and 4.893e+05, respectively, for the WA SEAHIRD model and 5.410e-07 and 7.477e+05, respectively, for the CO SEAHIRD model (see Figure 2). The curves for the other state variables in the SEAHIRD model, suggest that WA and CO had 42,598 and 55,295 symptomatic infected individuals (I + H), respectively, at their peak infection periods between March 15th and May 18th (see Figures S1 and S5). The number of asymptomatic infected cases (E + A), on the other hand, peaked at 302,310 and 401,103 individuals for WA and CO respectively during this period. In the SEAHIRD model, the peak number of hospitalizations occurred 24 and 36 days after March 15th for WA an CO, respectively. The actual peak in hospitalizations for these states during the modeling period, however, occurred 21 days and 39 days after March 15th, respectively. The probability of hospitalization given symptomatic infection was estimated as 0.0286 (see Figure 5A) and 0.0234 (see Figure 5B) for WA and CO, respectively. During the modeling period, CO started out as having fewer death cases than WA, but then overtook WA death counts by the end of the modeling period.

### 4.2 Effect of decreasing hospitalization on delaying bed shortage

Hospitalizing fewer COVID-19 patients delayed the time at which the bed capacity for COVID-19 patients was exceeded (Figure 3). This effect increased as fewer COVID-19 patients were hospitalized, according to the concave up shape of the graph in Figure 3. The curves for both WA and CO reach asymptotes; in other words, values of the hospitalization probability below which the bed cap will never be exceeded. This occurred around a probability of 0.0115 for WA and about 0.00551 for CO. The curve for CO was generally always higher than the curve for WA, reflecting how the peak infection period occurred later in CO than WA. However, there is a single point close to the WA asymptote where the CO and WA curves meet (Figure 3).

**Figure 3:**
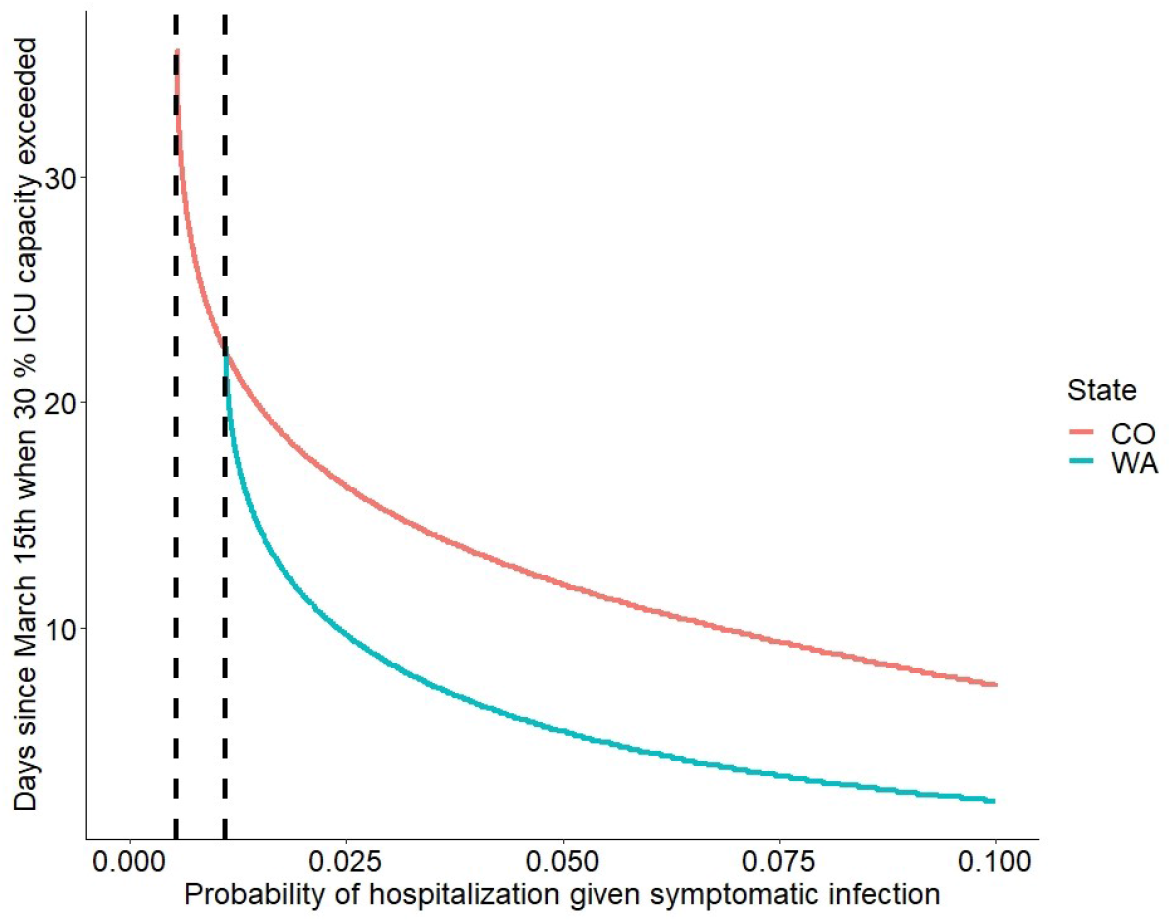
Dotted vertical lines represent asymptotes – the probability beyond which the ICU bed cap would not be exceeded from March 15th – May 18th. For both models *E*_0_ = 30\**I*_0_, *H*_0_ = 10\**h*_0_, and p = 0.86 and *χ* = 1.

### 4.3 Effect of increasing bed cap on delaying bed shortage

The graphs of Figure 4 show a concave down shape if the bed cap is less than 0.2. However, the graphs also inflect around values of 0.2 - 0.4 for the bed cap. Then, both curves approach an asymptote (dotted lines in Figure 4) as the bed cap increases. For WA, this asymptote occurred around a bed cap of 46 percent of the maximum. The asymptote for CO, on the other hand, occurred at a bed cap of about 60 percent of the maximum. For a given proportion of bed availability, bed caps were almost always exceeded later in CO than in WA.

**Figure 4:**
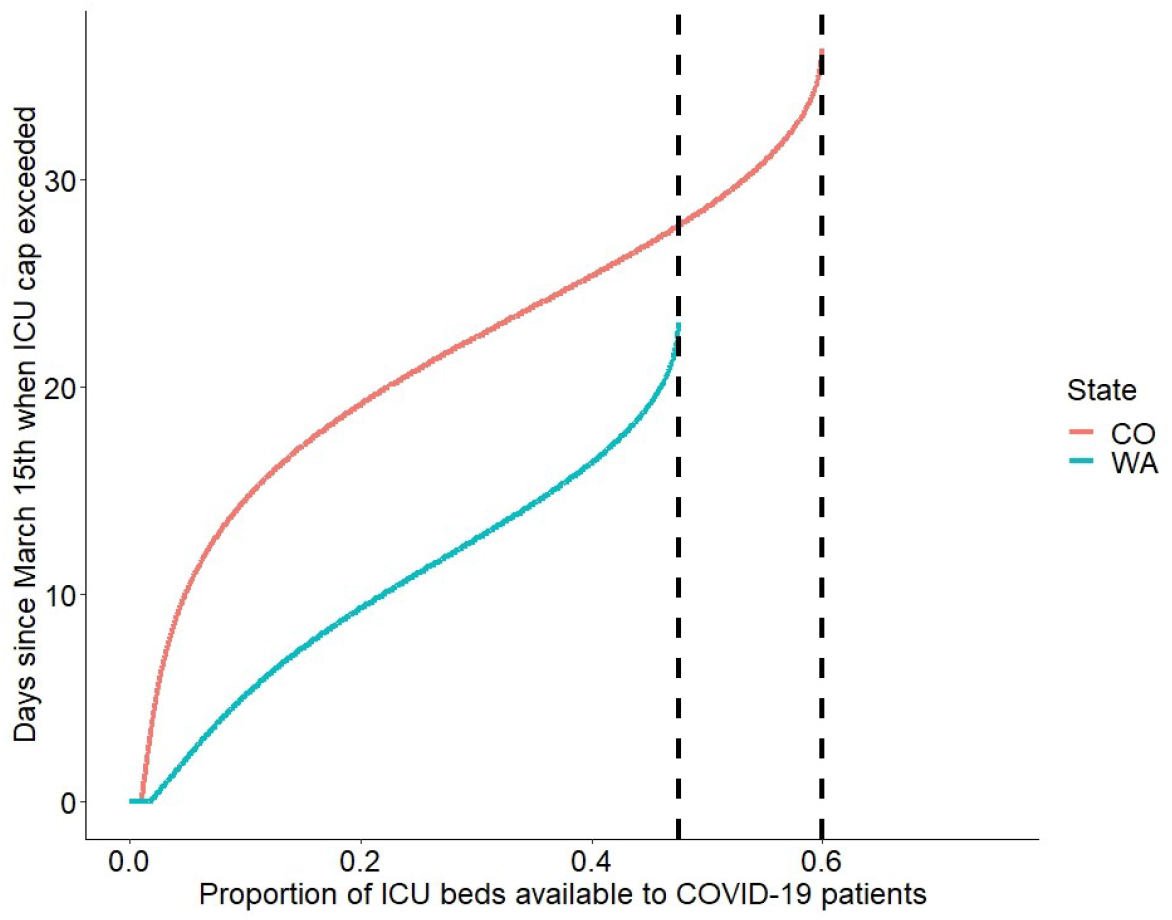
Dotted vertical lines represent asymptotes – proportion of ICU bed availability beyond which the ICU bed cap would not be exceeded from March 15th – May 18th. For both models *E*_0_ = 30\**I*_0_, *H*_0_ = 10\**h*_0_, and p = 0.86 and *χ* = 1.

### 4.4 SEAHIRD sensitivity to conditions and parameters

Once the SEAHIRD model was fit to each state’s data, we tested the local sensitivity of the SEAHIRD model to its initial conditions and *α*. We first used the local sensitivity analysis functions implemented in the FME package to identify the initial conditions with the largest influence on model output. For both states, the order from most sensitive to least sensitive parameters or conditions was: *S*_0_, *α, D*_0_, *E*_0_, *H*_0_ or *A*_0_, *I*_0_, and finally *R*_0_(see Figures S2, S3, S6, and S7). *S*_0_, *α*, and *D*_0_, are well-estimated either directly from data or from the model fitting, whereas the values of the remaining conditions were assumed. Thus, we took the two the most sensitive assumed initial conditions, *E*_0_ and *H*_0_, and altered their values, recording the effect on the SEAHIRD model’s predictions. For both states, altering *E*_0_ and *H*_0_ did not change the model’s prediction of when either state’s bed cap was exceeded by more than a day (see Tables S7 and S8). The bed cap was generally exceeded sooner when *E*_0_ = 40\**I*_0_ and *H*_0_ = 15\**h*_0_ but exceeded later when *E*_0_ = 20\**I*_0_ and *H*_0_ = 5\**h*_0_.

Two of the SEAHIRD model’s other primary assumptions are the values of *p* - the probability that an individual exposed to COVID-19 will develop an asymptomatic infection - and *χ* - the time it takes for symptomatic individuals to isolate themselves. Thus, we tested whether the SEAHIRD model’s predictions were robust to changing the value of *p*. We ran the SEAHIRD model with *p* set to 0.86 and 0.425, while keeping *E*_0_ = 30\**I*_0_, *H*_0_ = 10\**h*_0_. Lowering *p* to 0.425 noticeably affected the curves for both the WA and CO SEAHIRD models (see Figures S9 and S10). In the case of the WA SEAHIRD model with *p* = 0.425, the 30 percent bed capacity was expected to be exceeded about 3.7 days after March 15th. Decreasing hospitalization by 50 percent caused this capacity to never be exceeded while increasing bed capacity by 50 percent resulted in the bed capacity being exceeded 7.3 days after March 15th (see Table S9). The CO SEAHIRD model with *p* = 0.425 predicted that 30 percent ICU bed capacity of CO would be exceeded 14.4 days after March 15th. Decreasing the probability of hospitalization by 50 percent caused the 30 percent bed capacity to be reached 24.3 days after March 15th while increasing bed capacity by 50 percent resulted in an exceeded bed capacity 19.7 days after March 15th (see Table S9). The SEAHIRD model predictions under different values of *χ* are listed in Table S10.

## 5 Discussion

SARS-CoV-2 is unique to any other deadly virus humanity has ever faced. Its defining characteristics are that the majority of infected individuals do not develop symptoms and there is a significant period before symptoms could appear during which an individual can still spread the virus. This has led to massive levels of undocumented infection; some pre-print articles estimate that 19 million Americans have caught COVID-19, only a small fraction of which are laboratory confirmed cases [22]. We developed a novel system of differential equations to account for these asymptomatic and presymptomatic phases of COVID-19 infection, which happened to be similar to other models used to model COVID-19 spread [22]. This system was fit to data on COVID-19 mortality, the most reliable data available on COVID-19 spread, in two US states. These different states had different infection dynamics and different peak infection dates. Including infection peaks in the analysis was especially important because deterministic models can be especially misleading when fit with only pre-peak data [10]. We then compared the effects of decreasing the probability of hospitalization given symptomatic infection and increasing the proportion of ICU beds available to COVID-19 patients on delaying when the number of hospitalized COVID-19 patients exceeded the allotted ICU bed capacity. It is clear that, under the conditions simulated here, decreasing the probability of hospitalization has a larger effect on delaying an ICU bed shortage than comparably increasing the proportion of ICU beds available to COVID-19 patients. This finding was very robust; it occurred in the simulated conditions of both WA and CO under multiple sets of initial conditions and parameter values.

### 5.1 Decreasing hospitalization delays bed shortage more than increasing bed cap

At least two lines of evidence suggest that decreasing hospitalization of COVID-19 patients delays a bed shortage more than an equivalent increase in bed capacity. First, when we quantitatively compared times-till-bed-shortage across multiple sets of initial conditions and parameter values, scenarios with lower hospitalization of COVID-19 patients always exceeded ICU bed capacity at later dates – if at all – relative to scenarios with higher bed caps (see Tables S7 - S10). Furthermore, when we recorded the times-till-bed-shortage for thousands of combinations of hospitalization probability and bed cap values, we qualitatively observed decreasing the former to delay a bed shortage more than increasing the latter (Figures S4 and S8). This finding supports current CDC guidelines on COVID-19, which tell sick individuals to not go to hospitals unless they are experiencing severe COVID-19 symptoms [40].

Moderately changing either the bed cap or hospitalization probability rarely delayed a bed shortage by more than a few days from the baseline scenarios in our study. This seemingly small difference is arguably not clinically significant; however, COVID-19 regularly causes hundreds of deaths per day in the US (see https://coronavirus.jhu.edu/), making even small differences potentially lifesaving. This result is also promising because hospitalizing fewer patients is often more feasible for hospitals than increasing capacity. After all, increasing hospital capacity involves more than simply adding beds - extra staff, PPE, and medical equipment are also required for hospitals to care for more patients. Decreasing hospitalization and increasing hospital capacity are, of course, not mutually exclusive strategies. In fact, the saw-tooth pattern on our surface plots (Figures S4 and S8) suggests that multiple combinations of bed cap and hospitalization probability values can result in the bed cap never being exceeded for WA and CO. It is well-established, however, that some individuals are at relatively low risk of dying from COVID-19, especially individuals in younger age demographics [20] and individuals lacking comorbidities like obesity [38]. Thus, if such individuals can effectively isolate themselves at home, caring for fewer patients will leave more ICU beds open for seriously ill patients and therefore counter-intuitively save more lives. The danger is that it is impossible to predict with 100 percent certainty who will eventually develop deadly COVID-19 symptoms. Furthermore, policies to provide selective care could be enacted unfairly, increasing the already heavy toll of COVID-19 on marginalized groups in America [37]. Whatever policies are enacted to selectively care for COVID-19 patients, they must be evidence-based and equitable without exception.

### 5.2 SEAHIRD predictions are robust to initial conditions and parameter values

One of the most contentious parameters regarding COVID-19 disease spread is the probability of developing a symptomatic infection given COVID-19 exposure – denoted as *p* in this study. Early studies on COVID-19 suggested that this value could be fairly low – around 20 percent or less [16], [26]. Later studies suggested that this probability is likely higher - upwards of 50 percent [18],[19]. Thus, we tested whether the SEAHIRD model’s primary trend held true if we assumed both a low (0.425) and a high (0.86) value for *p*. Surprisingly, decreasing hospitalization by 50 percent always delayed a bed shortage more than increasing bed capacity by 50 percent and, in one case, avoided a bed shortage entirely (see Table S9). Changing the value of *p* had a large influence on the shape of the SEAHIRD model’s curves (see Figures S9 and S10). Thus, we suspect that changes in this parameter over time or could be responsible for the relatively poor fit between CO hospitalization data and the SEAHIRD model (see Figure 5B).

**Figure 5:**
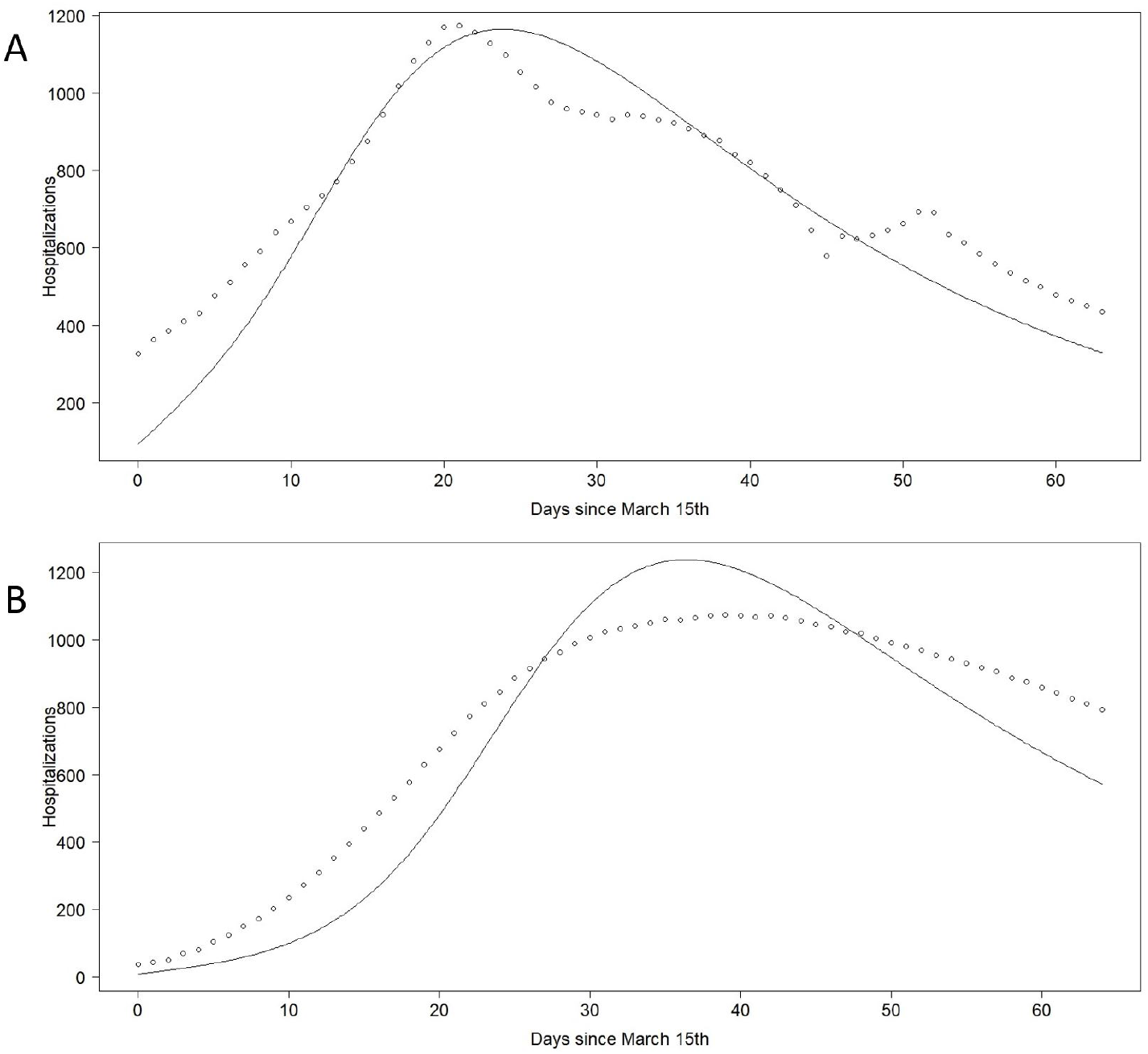
(A)Points are counts of hospitalized COVID-19 patients in all WA hospitals by day, corrected for underreporting by hospitals. Curve is H(t) function output from SEAHIRD model multiplied by the probability of hospitalization given symptomatic infection, which was numerically estimated as 0.0286. (B) Points are counts of hospitalized COVID-19 patients in all CO hospitals by day, corrected for underreporting by hospitals. Curve is H(t) function output from SEAHIRD model multiplied by the probability of hospitalization given symptomatic infection, which was estimated to be about 0.0234. For both (A) and (B), SEAHIRD initial conditions included *E*_0_ = 30\**I*_0_ and *H*_0_ = 10\**h*_0_ while p = 0.86 and *χ* = 1.

Our model assumed that all individuals self-isolate 24 hours after symptom onset, either by remaining home-bound or going to a hospital, which is optimistic relative to other COVID-19 models [1],[27],[33]. Nonetheless, we found that either decreasing this delay to 12 hours or increasing it to 48 hours led to the same general trend: increasing bed cap by 50 percent did not delay a bed shortage from being exceeded more than decreasing the hospitalization probability by 50 percent (see Table S10). Overall, the robustness of our main finding suggests it could hold for other populations with different infection dynamics.

### 5.3 Low effective population size for COVID-19 transmission in CO and WA

The effective population size of both the WA and CO SEAHIRD models was orders of magnitude lower than the actual population sizes of these states as estimated by the US census. This discrepancy is partly due to the lack of any lockdown effects in the SEAHIRD model. Thus, COVID-19 was restricted to spreading among only a subset of the WA and CO populations. The lack of lockdown effects is arguably SEAHIRD model’s most obviously violated assumption. WA lockdowns began on March 23rd 2020 [15], not long into the simulation, closely followed by CO on March 26th [23]. However, there is some pre-print work suggesting that US individuals were already relaxing physical distancing measures by mid-April, perhaps limiting the lockdown effect in our data [44]. Other phenomenon could have also contributed to the small effective population size in the SEAHIRD model. For example, there are well-documented cases of individuals unexposed to COVID-19 having antibodies that react to SARS-CoV-2 particles [32]. There’s also population-level evidence suggesting that certain pre-existing vaccines may have trained immune systems against COVID-19, although these vaccines are not common in the US [5],[12]. Nonetheless, it is possible that pre-existing and trained immunity may have removed individuals from the susceptible populations in CO and WA, explaining why COVID-19 only spread among a small subset of these populations in the SEAHIRD model. Incorporating lockdown effects, pre-existing immunity, and trained immunity into SEAHIRD-like models may improve their predictions. Understanding how these nuances affect hospitalizations will be incredibly important as the US tries to regain control of the COVID-19 pandemic.

## Data Availability

All of the data includedin this manuscript is available either in the supplemental section or in the provided links.

https://hgis.uw.edu/virus/

https://www.definitivehc.com/resources/covid-19-capacity-predictor

https://www.doh.wa.gov/Portals/1/Documents/2300/HospPatientData/YearEnd/Volume2018.xlsx

https://kdvr.com/wp-content/uploads/sites/11/2020/03/2018-ICU-Total-and-Licensed-Beds.pdf

https://www.census.gov/quickfacts/WA

https://www.census.gov/quickfacts/CO

