## Supplemental Figures and Tables for "Selectively caring for the most severe COVID-19 patients delays ICU bed shortages more than increasing hospital capacity"

| State | NoGrade | A+ | A | B | C | D | F | Ngrades | GPA |
|---|---|---|---|---|---|---|---|---|---|
| AK | 8 | 0 | 33 | 12 | 12 | 0 | 0 | 57 | 3.37 |
| AL | 8 | 0 | 0 | 53 | 4 | 0 | 0 | 57 | 2.93 |
| AR | 8 | 0 | 41 | 16 | 0 | 0 | 0 | 57 | 3.72 |
| AS | 40 | 0 | 0 | 0 | 7 | 3 | 14 | 24 | 0.71 |
| AZ | 8 | 24 | 0 | 33 | 0 | 0 | 0 | 57 | 3.84 |
| CA | 8 | 0 | 0 | 57 | 0 | 0 | 0 | 57 | 3.00 |
| CO | 8 | 0 | 40 | 17 | 0 | 0 | 0 | 57 | 3.70 |
| CT | 8 | 0 | 33 | 18 | 6 | 0 | 0 | 57 | 3.47 |
| DC | 8 | 7 | 41 | 9 | 0 | 0 | 0 | 57 | 3.96 |
| DE | 8 | 0 | 19 | 24 | 14 | 0 | 0 | 57 | 3.09 |
| FL | 8 | 7 | 33 | 3 | 14 | 0 | 0 | 57 | 3.58 |
| GA | 8 | 7 | 46 | 4 | 0 | 0 | 0 | 57 | 4.05 |
| GU | 40 | 0 | 0 | 0 | 10 | 6 | 8 | 24 | 1.08 |
| HI | 8 | 0 | 33 | 19 | 5 | 0 | 0 | 57 | 3.49 |
| IA | 8 | 12 | 21 | 10 | 14 | 0 | 0 | 57 | 3.54 |
| ID | 8 | 0 | 33 | 12 | 12 | 0 | 0 | 57 | 3.37 |
| IL | 8 | 0 | 57 | 0 | 0 | 0 | 0 | 57 | 4.00 |
| IN | 8 | 19 | 1 | 37 | 0 | 0 | 0 | 57 | 3.68 |
| KS | 8 | 0 | 20 | 33 | 4 | 0 | 0 | 57 | 3.28 |
| KY | 8 | 7 | 50 | 0 | 0 | 0 | 0 | 57 | 4.12 |
| LA | 8 | 0 | 32 | 25 | 0 | 0 | 0 | 57 | 3.56 |
| MA | 8 | 7 | 13 | 37 | 0 | 0 | 0 | 57 | 3.47 |
| MD | 8 | 0 | 43 | 7 | 0 | 7 | 0 | 57 | 3.51 |
| ME | 8 | 0 | 33 | 24 | 0 | 0 | 0 | 57 | 3.58 |
| MI | 8 | 24 | 0 | 29 | 0 | 4 | 0 | 57 | 3.70 |
| MN | 8 | 6 | 51 | 0 | 0 | 0 | 0 | 57 | 4.11 |
| MO | 8 | 0 | 26 | 17 | 14 | 0 | 0 | 57 | 3.21 |
| MP | 40 | 0 | 0 | 0 | 0 | 24 | 0 | 24 | 1.00 |
| MS | 8 | 0 | 31 | 26 | 0 | 0 | 0 | 57 | 3.54 |
| MT | 8 | 0 | 29 | 7 | 21 | 0 | 0 | 57 | 3.14 |
| NC | 8 | 0 | 40 | 17 | 0 | 0 | 0 | 57 | 3.70 |
| ND | 8 | 0 | 0 | 33 | 24 | 0 | 0 | 57 | 2.58 |
| NE | 8 | 0 | 33 | 2 | 5 | 17 | 0 | 57 | 2.89 |
| NH | 8 | 0 | 0 | 46 | 11 | 0 | 0 | 57 | 2.81 |
| NJ | 8 | 21 | 32 | 4 | 0 | 0 | 0 | 57 | 4.30 |
| NM | 8 | 0 | 33 | 0 | 24 | 0 | 0 | 57 | 3.16 |
| NV | 8 | 0 | 33 | 6 | 3 | 11 | 4 | 57 | 2.93 |
| NY | 8 | 0 | 38 | 19 | 0 | 0 | 0 | 57 | 3.67 |
| OH | 8 | 0 | 0 | 55 | 0 | 2 | 0 | 57 | 2.93 |
| OK | 8 | 12 | 19 | 26 | 0 | 0 | 0 | 57 | 3.75 |
| OR | 8 | 16 | 37 | 4 | 0 | 0 | 0 | 57 | 4.21 |
| PA | 8 | 0 | 33 | 11 | 13 | 0 | 0 | 57 | 3.35 |
| PR | 7 | 0 | 33 | 3 | 6 | 15 | 0 | 57 | 2.95 |
| RI | 8 | 24 | 0 | 33 | 0 | 0 | 0 | 57 | 3.84 |
| SC | 8 | 0 | 8 | 49 | 0 | 0 | 0 | 57 | 3.14 |
| SD | 8 | 0 | 33 | 7 | 17 | 0 | 0 | 57 | 3.28 |
| TN | 8 | 0 | 39 | 18 | 0 | 0 | 0 | 57 | 3.68 |
| TX | 8 | 0 | 40 | 17 | 0 | 0 | 0 | 57 | 3.70 |
| UT | 8 | 0 | 33 | 12 | 12 | 0 | 0 | 57 | 3.37 |
| VA | 8 | 15 | 33 | 9 | 0 | 0 | 0 | 57 | 4.11 |
| VI | 40 | 0 | 0 | 0 | 20 | 4 | 0 | 24 | 1.83 |
| VT | 8 | 0 | 33 | 14 | 10 | 0 | 0 | 57 | 3.40 |
| WA | 8 | 0 | 25 | 17 | 15 | 0 | 0 | 57 | 3.18 |
| WI | 8 | 17 | 40 | 0 | 0 | 0 | 0 | 57 | 4.30 |
| WV | 8 | 0 | 0 | 43 | 14 | 0 | 0 | 57 | 2.75 |
| WY | 8 | 0 | 33 | 7 | 17 | 0 | 0 | 57 | 3.28 |

Table S1: evaluation of COVID-19 data tracking regularity during period of March 15th – May 18th. Data is from COVID Tracking Project, a volunteer organization hosted by *The Atlantic* dedicated to collecting and publishing COVID-19 testing data in the US (https://covidtracking.com/data/download). The grade measures only the consistency of reporting, not necessarily the quality of the data. Ngrades stands for number of grades, the sum of columns A+ through F. The NoGrade column gives the number of days where a state did not get a grade. The states used in this study are highlighted in green. GPA was calculated as (A+*5 + A*4 + B*3 + C*2 + D*1 + F*0)/ Ngrades where A+, A, B, C, D, and F represent the number of times a state got each of those respective grades.

Table S2: Sources of epidemiological COVID-19 data

| Data type | Source |
|---|---|
| Counts of deaths from COVID-19 | https://hgis.uw.edu/virus/ |
| Counts of hospitalizations from COIVD-19 | https://www.definitivehc.com/resources/covid-19-capacity-predictor |
| Number of ICU beds in WA | https://www.doh.wa.gov/Portals/1/Documents/2300/HospPatientData/YearEnd/Volume2018.xlsx |
| Number of ICU beds in CO | https://kdvr.com/wp-content/uploads/sites/11/2020/03/2018-ICU-Total-and-Licensed-Beds.pdf |
| WA population size | https://www.census.gov/quickfacts/WA |
| CO population size | https://www.census.gov/quickfacts/CO |

Table S3: hospital reporting frequency in WA from May 9th – May 15th

| Date | Hospitals Reported | Percent of all acute care hospitals reported |
|---|---|---|
| 5/9/2020 | 50 | 54.3 |
| 5/10/2020 | 48 | 52.2 |
| 5/11/2020 | 82 | 89.1 |
| 5/12/2020 | 80 | 87.0 |
| 5/13/2020 | 76 | 82.6 |
| 5/14/2020 | 75 | 81.5 |
| 5/15/2020 | 51 | 55.4 |

Table S3: Data used to correct for underreporting by WA hospitals. Data is no longer available on WA DOH website. There are 92 acute care hospitals in WA.

Table S4: hospitalization reporting frequency by day for CO hospitals

| Date | Percentage of hospitals reported |
|---|---|
| July 3 | 76 |
| July 4 | 65 |
| July 5 | 56 |
| July6 | 76 |
| July7 | 86 |
| July 8 | 86 |
| July 9 | 83 |

Table S4: data used to correct for underreporting in CO hospitals. Assumed to be representative of the period March 15th – May 18th, especially since US is still in the midst of the pandemic. Data is no longer available on the CO DOH website.

Table S5: SEAHIRD model parameters and initial conditions

| Parameter | Definition | Value | References |
|---|---|---|---|
| α | Contact rate between susceptible and infected individuals | Determined by optimization | NA |
| β | Rate constant for incubation of COVID-19 (inverse of incubation period) | 1/3.60 days | (Li *et al.* 2020) |
| ζ | recovery time for infection | 24.7 days | (Verity *et al.* 2020) |
| χ | time until self-isolation once symptoms appear | 0.5, 1, or 2 days | NA |
| p | probability of developing an asymptomatic infection, given exposure to COVID-19 | 0.86 or 0.425 | (Li *et al.* 2020; Lavezzo et al. 2020) |
| q | probability of dying, given a symptomatic infection with COVID-19 | 0.986 | (Wu *et al.* 2020) |
| S0 | The number of susceptible individuals in Washington on March 15th | N – E0 – A0 – H0 – I0 – R0 – D0 | https://www.census.gov/quickfacts/WA https://www.census.gov/quickfacts/CO |
| E0 | The number of exposed individuals in Washington on March 15th | $40I_0$, $30I_0$, or $20I_0$ | NA |
| A0 | The number of asymptomatic, infected individuals in Washington on March 15th | $4*I_0$ | NA |
| H0 | The number of symptomatic, infected, and isolated individuals in Washington on March 15$^{th}$ | $15h_0$, $10h_0$, or $5h_0$ | NA |
| I0 | The number of symptomatic, infected, and not-isolated individuals in Washington on March 15th | $1.79C_0 – D_0$ | (Lachmann 2020) |
| R0 | The number of individuals that have recovered from a symptomatic or asymptomatic infection in Washington on March 15$^{th}$ | 0 | NA |
| D0 | The number of individuals that died from COVID-19 infection on March 15$^{th}$ | Determined directly from data | https://hgis.uw.edu/virus/ |

Table S6: State variable definitions

| State variable | Definition |
|---|---|
| S | Susceptible individuals |
| E | Exposed individuals |
| A | Individuals that are infected but not symptomatic |
| I | Individuals that are infected and symptomatic |
| H | Individuals that are infected, symptomatic, and isolated (i.e. either hospitalized or homebound) |
| R | Individuals that recovered from either a symptomatic or asymptomatic infection |
| D | Individuals that died from COVID-19 infection |

Table S7: Sensitivity analysis results for WA

| | | E0 | | |
|---|---|---|---|---|
| | | 40*I0 | 30*I0 | 20*I0 |
| H0 | 15*h0 | 8.6/15.8/12.3 | 9.1/15.7/12.4 | 9.5/15.5/12.6 |
| | 10*h0 | 8.9/15.5/12.2 | 8.3/15.7/11.9 | 9.6/15.2/12.4 |
| | 5*h0 | 9.1/15.2/12.1 | 9.4/15/12.2 | 9.7/14.9/12.3 |

Table S7: All numbers are days from March 15$^{th}$ when 30 % of ICU bed capacity is exceeded. In each cell, the first number gives days until bed cap is exceeded if no action is taken, second number gives days until bed cap is exceeded if probability of hospitalization given symptomatic infection is decreased by 50 %, third number gives days until bed cap is exceeded if ICU bed cap is increased by 50 %.

Table S8: Sensitivity analysis results for CO

| | | E0 | | |
|---|---|---|---|---|
| | | 40*I0 | 30*I0 | 20*I0 |
| H0 | 15*h0 | 16.1/21.6/19.2 | 16.5/21.7/19.4 | 17/21.8/19.6 |
| | 10*h0 | 16.1/21.6/19.1 | 16.5/21.7/19.4 | 17/21.7/19.6 |
| | 5*h0 | 16.2/21.6/19.1 | 16.5/21.6/19.3 | 17/21.7/19.6 |

Table S8: All numbers are days from March 15$^{th}$ when 30 % of ICU bed capacity is exceeded. In each cell, the first number gives days until bed cap is exceeded if no action is taken, second number gives days until bed cap is exceeded if probability of hospitalization given symptomatic infection is decreased by 50 %, third number gives days until bed cap is exceeded if ICU bed cap is increased by 50 %.

Table S9: Effect of the probability of symptomatic infection given COVID-19 exposure on SEAHIRD model predictions

| p value | WA SEAHIRD model predictions (days till bed cap exceeded) | CO SEAHIRD model predictions (days till bed cap exceeded) |
|---|---|---|
| 0.86 | 8.3/15.7/11.9 | 16.5/21.7/19.4 |
| 0.425 | 3.7/NA/7.3 | 14.4/24.3/19.7 |

Table S9: All numbers are counts of days past March 15$^{th}$, when the SEAHIRD model simulations began. In each cell, the first number gives days until bed cap is exceeded if no action is taken, second number gives days until bed cap is exceeded if probability of hospitalization given symptomatic infection is decreased by 50 %, third number gives days until bed cap is exceeded if bed cap is increased by 50 %. While the value of p changes between cells, all other parameters and conditions were held at their baseline values. NA means that the bed cap was not exceeded during the modeling period.

Table S10: Effect of time till self-isolation on SEAHIRD model predictions

| $\chi$ value | WA SEAHIRD model predictions (days till bed cap exceeded) | CO SEAHIRD model predictions (days till bed cap exceeded) |
|---|---|---|
| 48 hours | 8.7/15.6/12.1 | 16.7/21.7/19.5 |
| 24 hours | 8.3/15.7/11.9 | 16.5/21.7/19.4 |
| 12 hours | 8.1/15.7/11.9 | 16.4/21.6/19.3 |

Table S10: All numbers are counts of days past March 15$^{th}$, when the SEAHIRD model simulations began. In each cell, the first number gives days until bed cap is exceeded if no action is taken, second number gives days until bed cap is exceeded if probability of hospitalization given symptomatic infection is decreased by 50 %, third number gives days until bed cap is exceeded if ICU bed cap is increased by 50 %. While the value of $\chi$ changes between cells, all other parameters and conditions were held at their baseline values.

Figure S1: all curves for the baseline WA SEAHIRD model state variables

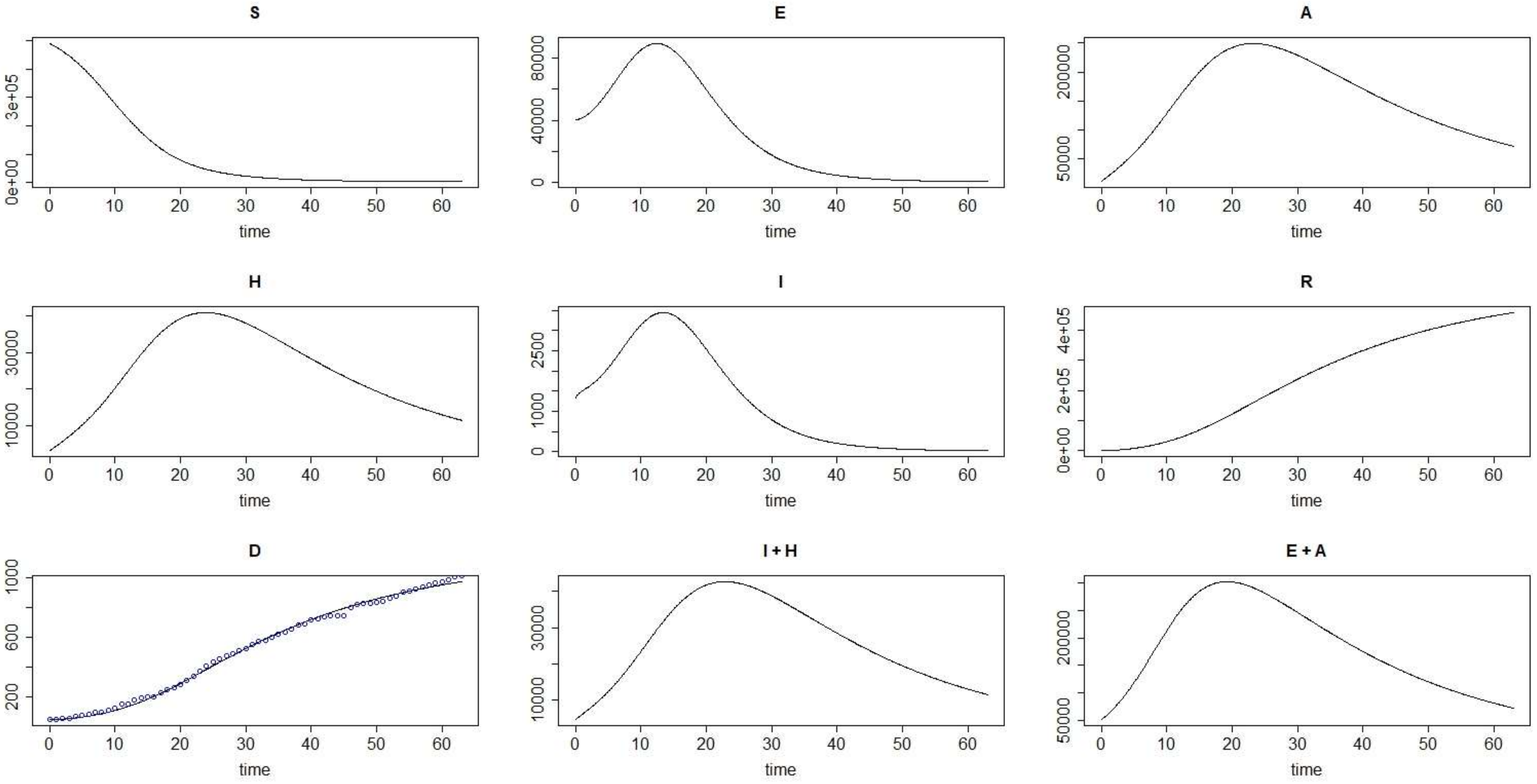


Figure S1: State variables, given at the top of each graph are defined in Table 1. The points in curve D are actual WA mortality data while the line is the SEAHIRD model fit. All y-axes are counts of individual cases and all x-axes have units of day. SEAHIRD initial conditions included $E_0 = 30*I_0$ and $H_0 = 10*h_0$ while $p = 0.86$ and $\chi = 1$. Plots are labeled according to the SEAHIRD model compartment they correspond to (see Table 2). I + H represents the total number of symptomatic infections while E + A represents the total number of asymptomatic infections.

Figure S2: Sensitivity functions for WA SEAHIRD model

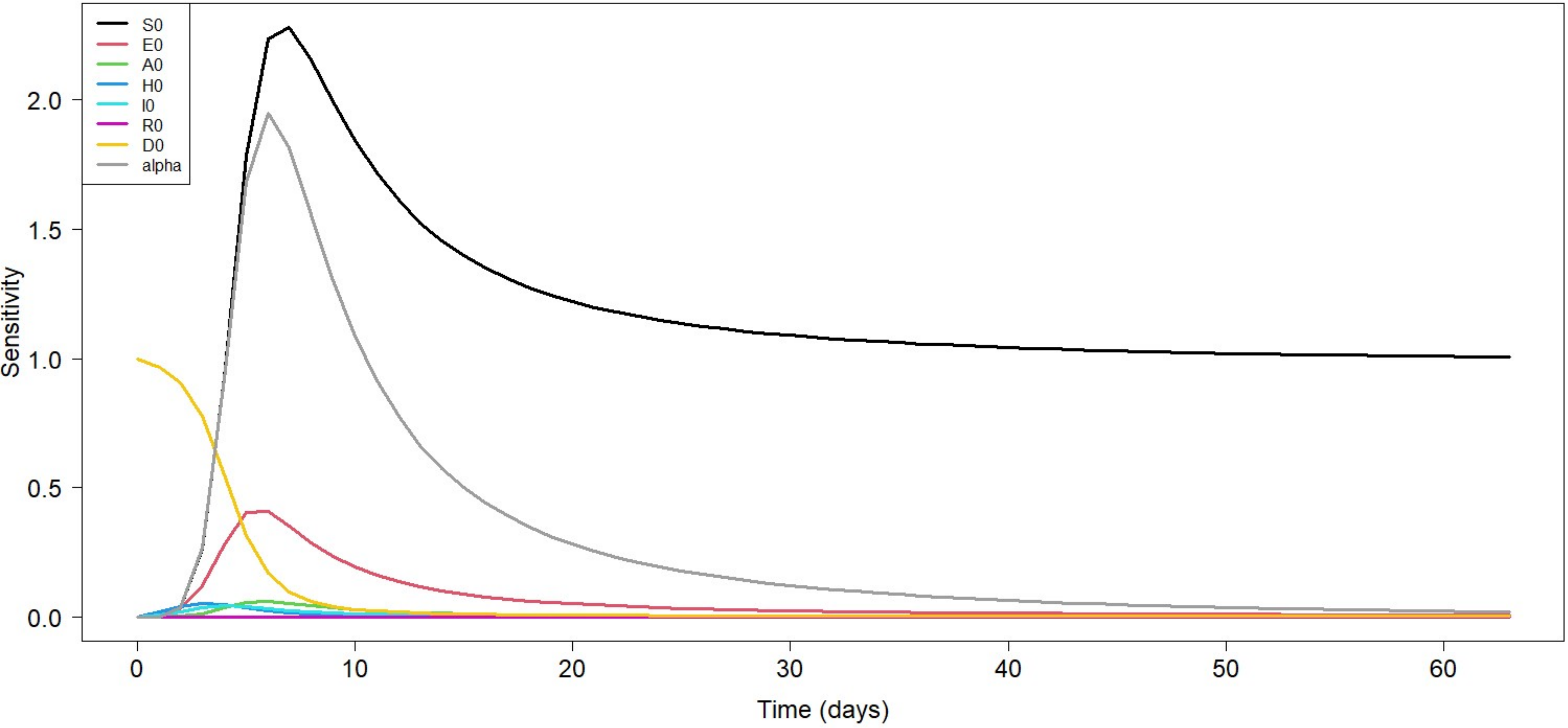


Figure S2: SEAHIRD initial conditions included $E_0 = 30*I_0$ and $H_0 = 10*h_0$ while $p = 0.86$ and $\chi= 1$. This plot was generated with the FME package in R.

Figure S3: Summary of sensitivity functions for WA SEAHIRD model

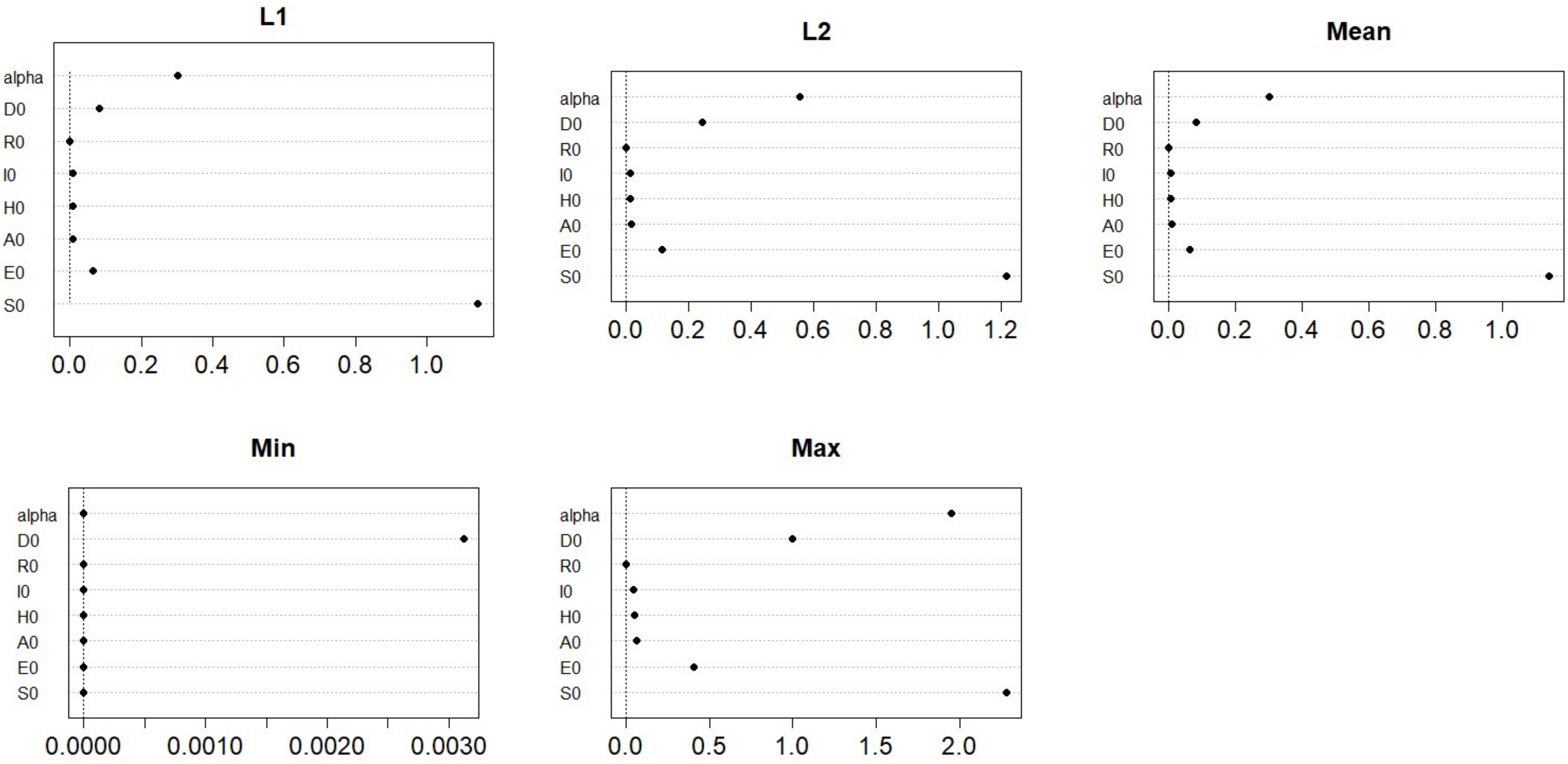


Figure S3: SEAHIRD initial conditions included $E_0 = 30*I_0$ and $H_0 = 10*h_0$ while $p = 0.86$ and $\chi = 1$. These plots were generated with the FME package in R.

Figure S4: Effect of simultaneously changing the probability of hospitalization given symptomatic infection and the proportion of ICU beds available to COVID-19 patients in WA.

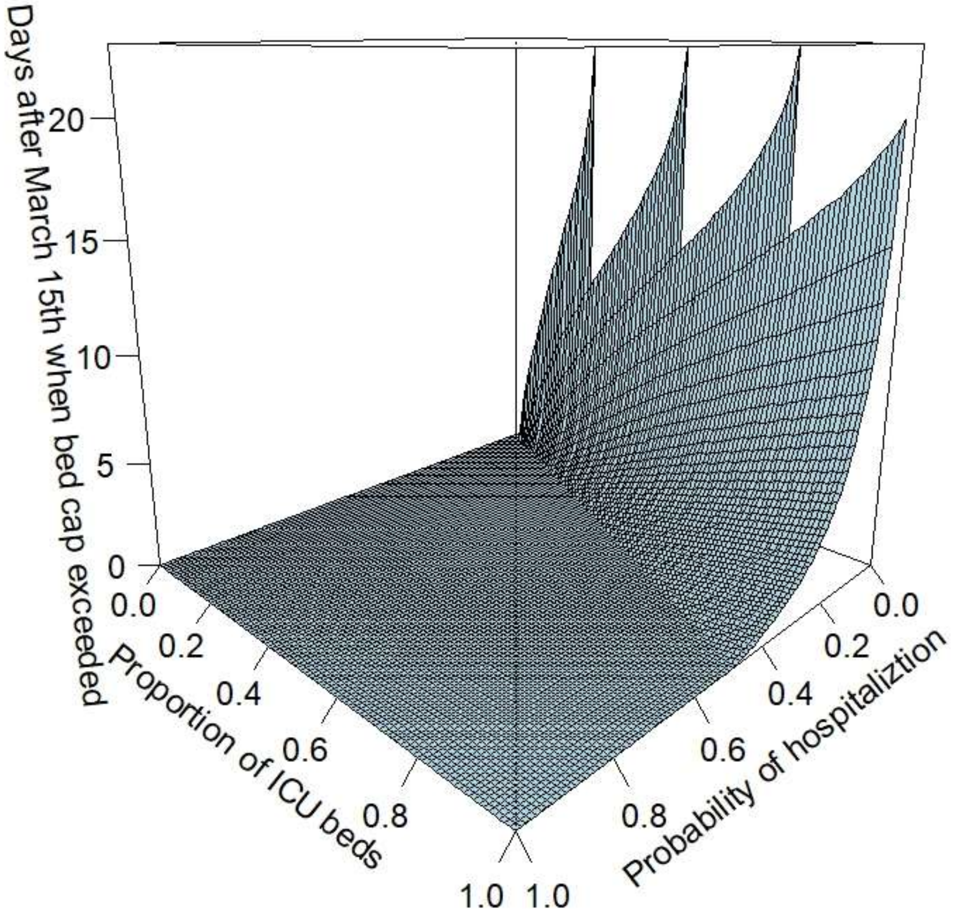


Figure S4: surface was generated by plugging in 10,201 combinations of values for the probability of hospitalization given symptomatic infection and the proportion of ICU beds available to COVID-19 patients into the WA SEAHIRD model and seeing when hospitalized COVID-19 cases exceeded the proportion of ICU beds available * total ICU beds in state. If a particular combination of these values caused the threshold number of beds to not be exceeded, then no point surface was generated at that point. In the case of this model, $E_0 = 30*I_0$, $H_0 = 10*h_0$, and $p = 0.86$ and $\chi = 1$.

Figure S5: all curves for baseline CO SEAHIRD model

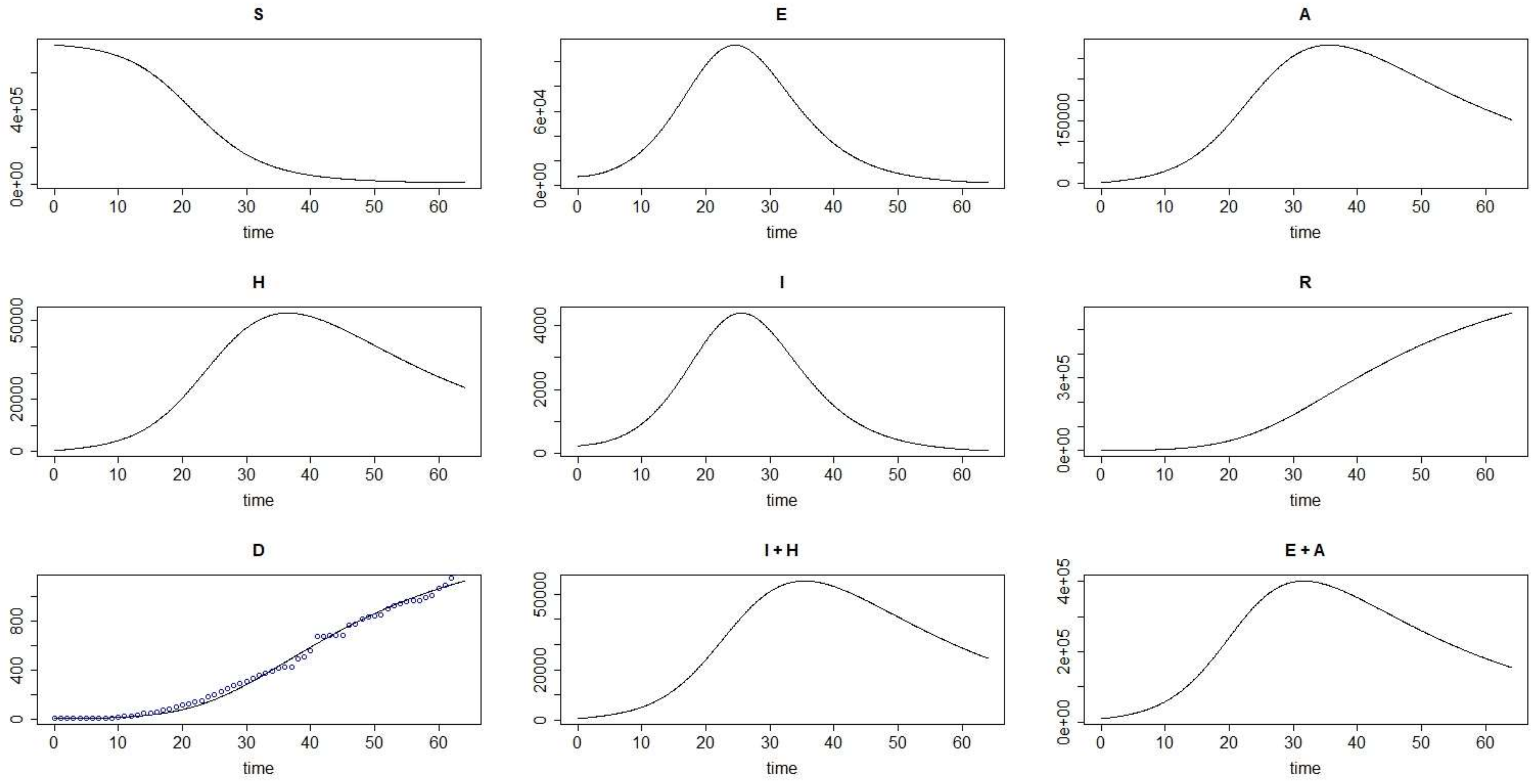


Figure S5: State variables given at the top of each graph are defined in Table 1. The points in curve D are actual CO mortality data while the line is the SEAHIRD model fit. All y-axes are counts of individual cases and all x-axes have units of days. SEAHIRD initial conditions included $E_0 = 30*I_0$ and $H_0 = 10*h_0$ while p = 0.86 and $\chi = 1$. Plots are labeled according to the SEAHIRD model compartment they correspond to (see Table S6). I + H represents the total number of symptomatic infections while E + A represents the total number of asymptomatic infections.

Figure S6: Summary of sensitivity functions for CO SEAHIRD model

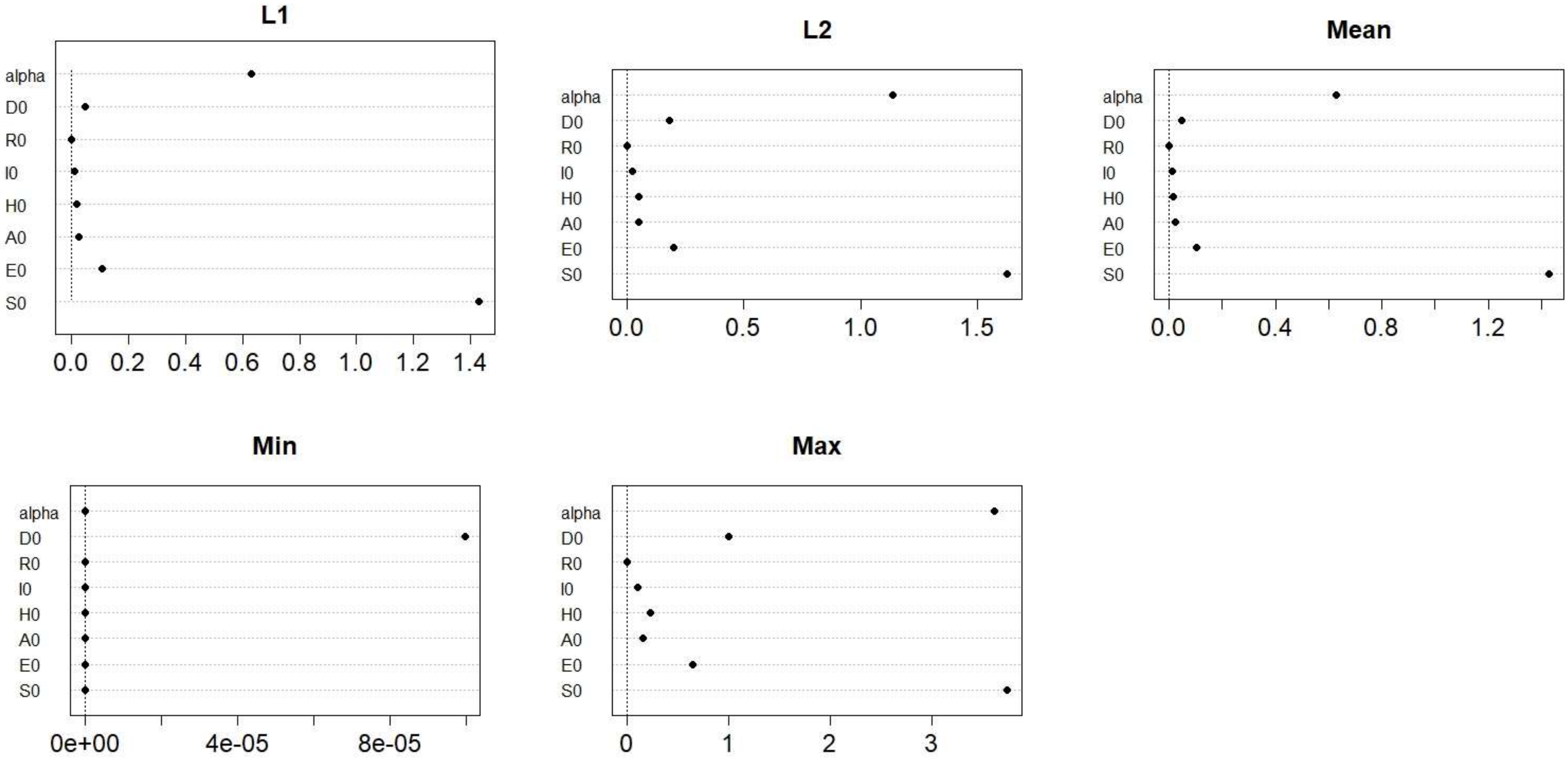


Figure S6: SEAHIRD initial conditions included $E_0 = 30*I_0$ and $H_0 = 10*h_0$ while $p = 0.86$ and $\chi = 1$. These plots were generated with the FME package in R.

Figure S7: Sensitivity functions for baseline CO SEAHIRD model

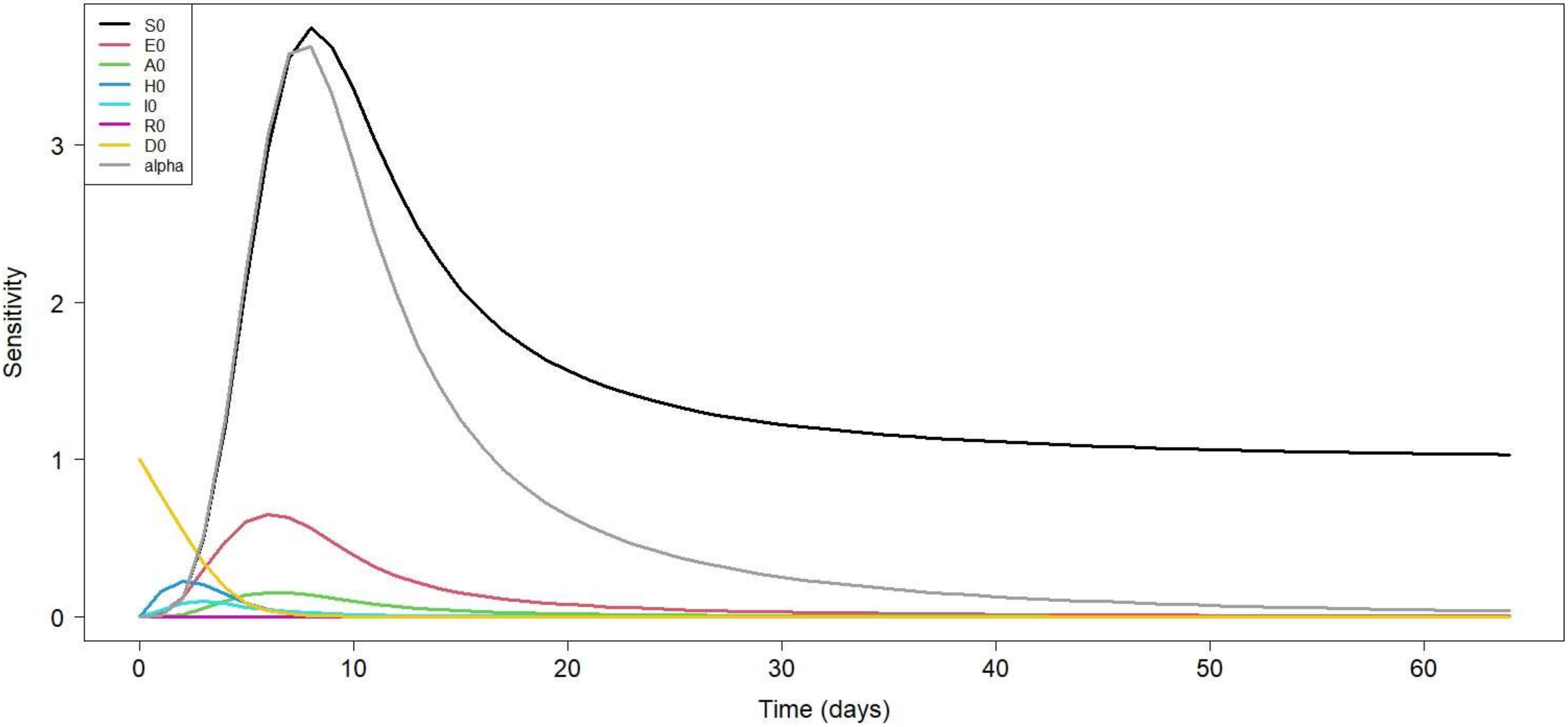


Figure S7: SEAHIRD initial conditions included $E_0 = 30*I_0$ and $H_0 = 10*h_0$ while $p = 0.86$ and $\chi = 1$. This plot was generated with the FME package in R.

Figure S8: Effect of simultaneously changing the probability of hospitalization given symptomatic infection and the proportion of ICU beds available to COVID-19 patients in CO.

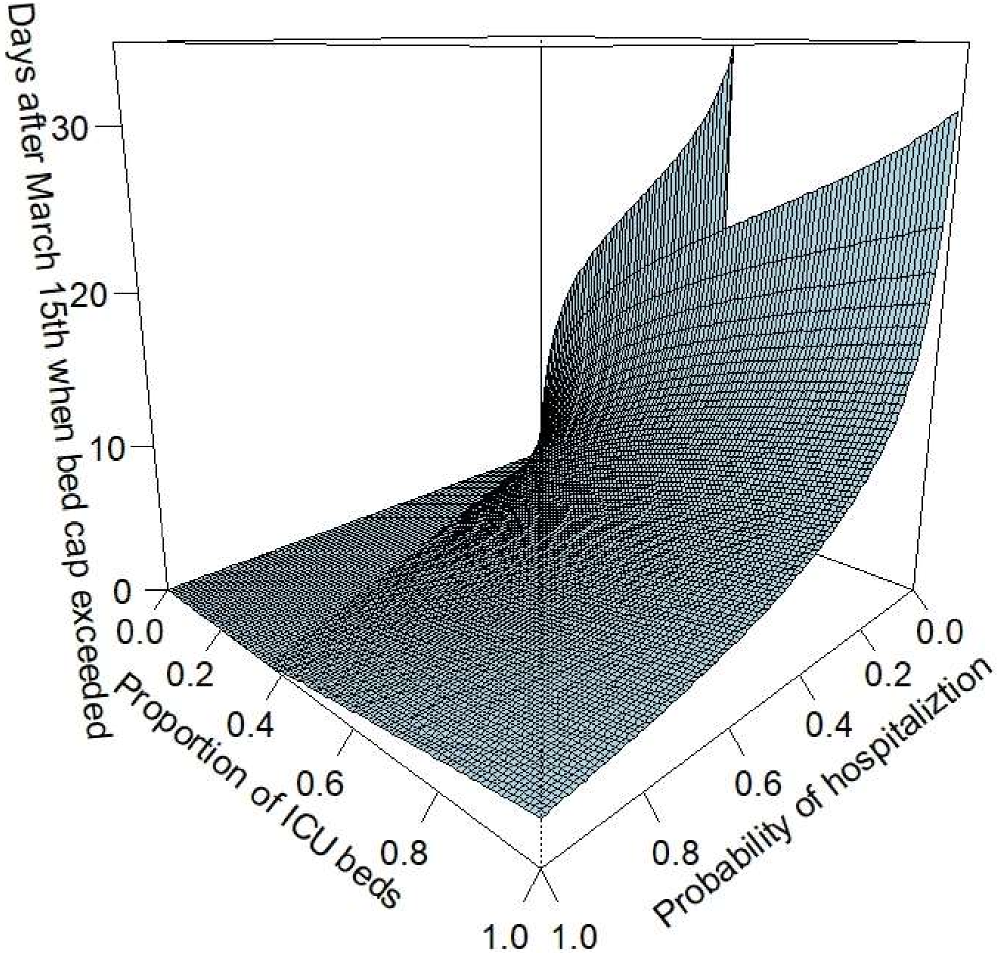


Figure S8: Surface was generated by plugging in 10,201 combinations of values for the probability of hospitalization given symptomatic infection and the proportion of ICU beds available to COVID-19 patients into the CO SEAHIRD model and seeing when hospitalized COVID-19 cases exceeded the proportion of ICU beds available * total ICU beds in state. If a particular combination of these values caused the threshold number of beds to not be exceeded, then no point surface was generated at that point. In the case of this model, $E_0 = 30*I_0$, $H_0 = 10*h_0$, and $p = 0.86$ and $\chi = 1$.

Figure S9: SEAHIRD model curves for WA with lower probability of developing symptoms given COVID-19 exposure relative to baseline model

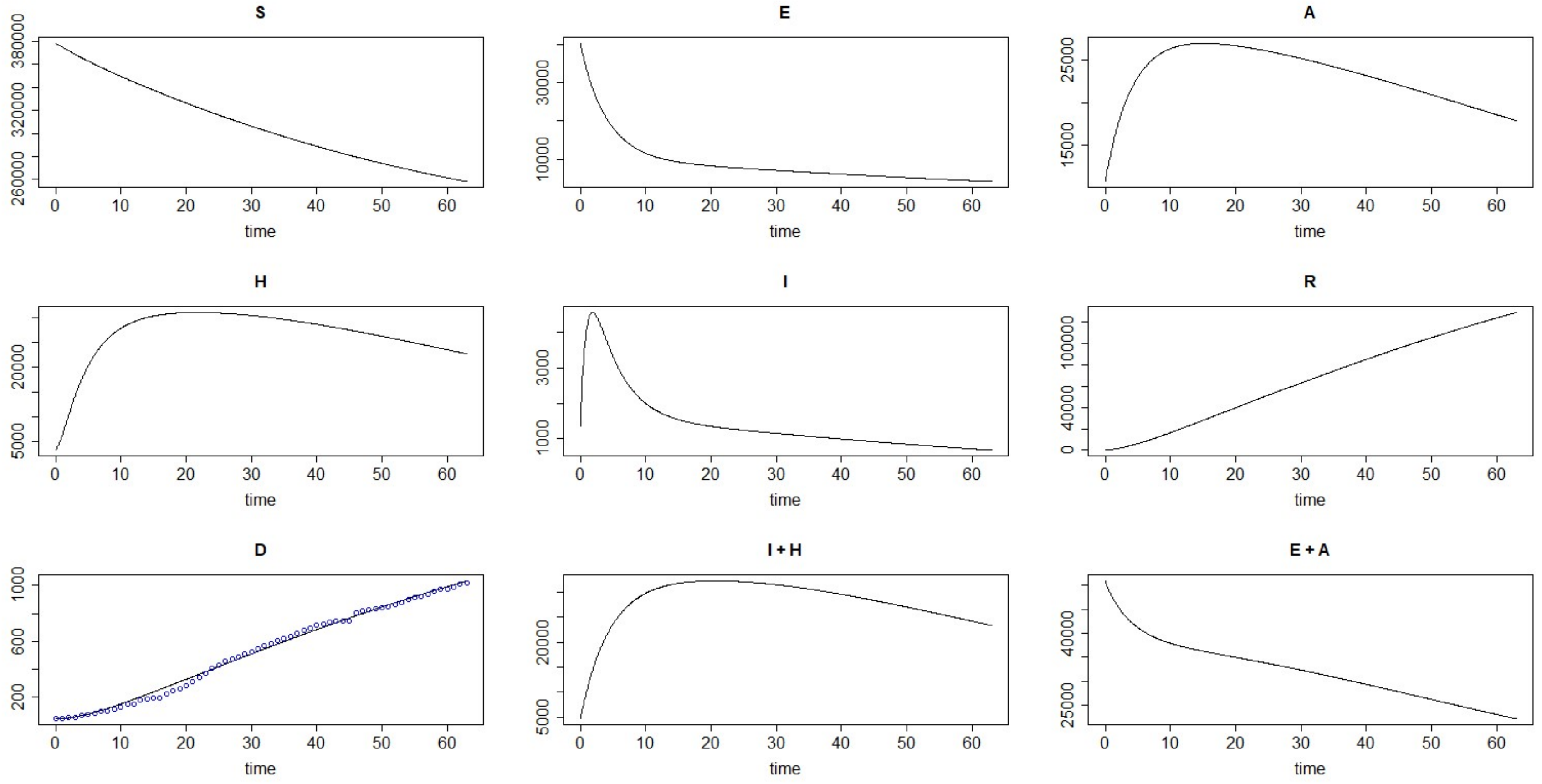


Figure S9: SEAHIRD initial conditions included $E_0 = 30*I_0$ and $H_0 = 10*h_0$, while $p = 0.425$ and $\chi = 1$. Plots are labeled according to the SEAHIRD model compartment they correspond to (see Table S6). I + H represents the total number of symptomatic infections while E + A represents the total number of asymptomatic infections.

Figure S10: SEAHIRD model curves for CO with lower probability of developing symptoms given COVID-19 exposure relative to baseline model

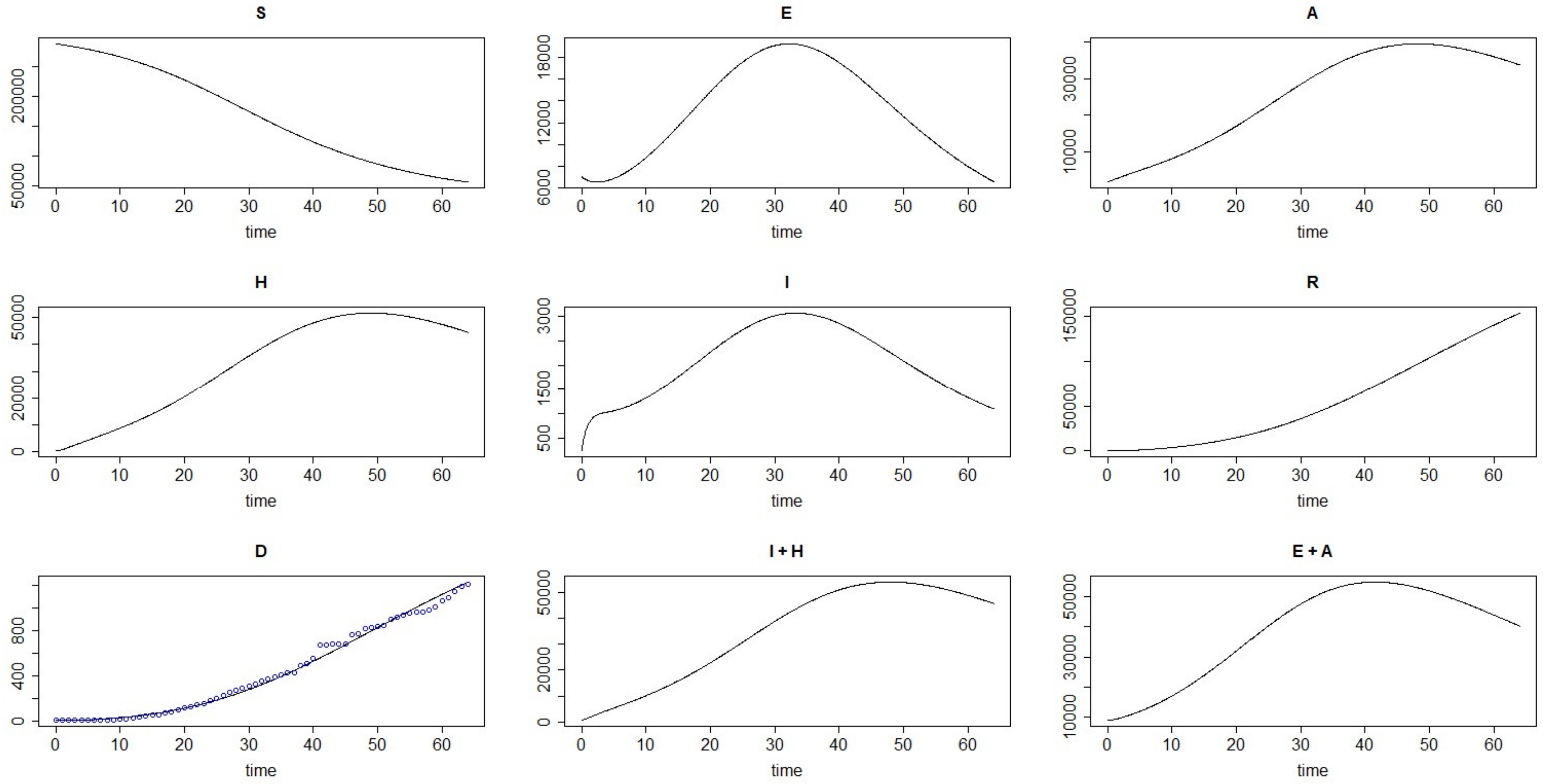


Figure S10: SEAHIRD initial conditions included $E_0 = 30*I_0$ and $H_0 = 10*h_0$, while $p = 0.425$ and $\chi = 1$. Plots are labeled according to the SEAHIRD model compartment they correspond to (see Table S6). I + H represents the total number of symptomatic infections while E + A represents the total number of asymptomatic infections.
